# Integrating Ethnography and Structural Equation Modelling to Assess Brucellosis Knowledge, Attitude, and Practices among Pastoralist Communities in Kenya

**DOI:** 10.1101/2025.10.17.25338227

**Authors:** Dismas C.O. Oketch, Samuel Waiguru Muriuki, Dalmas Omia, Bonface Osindi, Ruth Njoroge, Justice Amenyo Kessie, Boku Bodha, John Gachohi, Scott L Nuismer, Nazaria Wanja Nyaga, John Mwaniki Njeru, Samoel Ashimosi Khamadi, Humphrey Kariuki Njaanake, Eunice Kamaara, Walter Jaoko, M. Kariuki Njenga, Eric Osoro

**Author notes:** Corresponding author: Dismas Oketch [ ]] Washington State University Global Health-Kenya, One Padmore Place, George Padmore Road, Nairobi. P.O. Box 72938-00200, Nairobi, Kenya Telephone: Office Line: www.globalhealth.wsu.edu.

## Abstract

**Background:** Brucellosis is a neglected zoonotic disease endemic in pastoralist communities. It is driven by close human-animal interactions and culturally embedded practices such as consuming raw milk and blood, handling aborted materials, and skins and hides, and limited access to veterinary and healthcare services. Understanding how knowledge, practices, and cultural beliefs intersect structurally is essential for designing context specific One Health interventions for effective disease control.

**Methods:** We employed a mixed-methods approach to explore brucellosis-related knowledge, attitudes, and practices among pastoralist communities in Kajiado and Marsabit counties, in Kenya. Subsequently, we used partial least squares structural equation modeling (PSL-SEM) to estimate the direct and indirect effect of the latent variables (K, A, P) on the brucellosis transmission pathway.

**Results:** Brucellosis knowledge, attitude, and practices were markedly poor in both counties. Pastoralists in Marsabit demonstrated significantly better knowledge (54% vs 36%, p<0.001), and attitude (45% vs 23%, p<0.001) but worse practice scores (26% vs 52%, p<0.001), compared to Kajiado. Risky practices, such as raw milk consumption and unsafe disposal of animal birthing products, were widespread. PLS-SEM analysis showed a strong indirect effect of knowledge on practices mediated through attitudes: (Knowledge → Attitude: β = 0.921, p<0.001; Attitude → Practice: β = 0.580, p< 0.001), with minimal direct impact (Knowledge → Practice: β = 0.188). Cultural beliefs, such as resistance to boiling milk or using colostrum for healing – persisted regardless of awareness levels.

**Discussion:** This study reinforces the observation that knowledge alone is insufficient for behavior change. Cultural norms, traditional knowledge systems, and limited health infrastructure play a critical role in sustaining risk behaviors.

**Conclusions:** Effective brucellosis control must integrate culturally grounded, community-specific strategies that strengthen attitudes and promote safer practices within a One Health framework.

**KEY MESSAGES:** 

**What is already known on this topic:**

- Brucellosis transmission is shaped by deeply embedded cultural practices and low levels of disease awareness among pastoralist communities.
- Persistent risky sanitary and dietary practices in pastoralist communities is driven by culture and survival necessity trade-offs.
- Previous studies on brucellosis transmission among pastoralist communities often rely on quantitative surveys that miss cultural and contextual nuances.

**What this study adds:**

- Our study provides clear understanding of how knowledge, attitudes, and practices (KAP) interact structurally within pastoralist communities.
- The study bridges the gap between qualitative cultural insights and quantitative modeling to better inform zoonotic disease control interventions.
- The study demonstrates that combining ethnography with Structural Equation Modelling (SEM) provides a richer, more nuanced understanding of KAP dynamics around brucellosis.
- It reveals that cultural beliefs and attitudes significantly mediate the structural interaction between knowledge and practices, and confirms that knowledge alone isn’t adequate to influence safe dietary and sanitary practices.
- The SEM model identified key latent variables such as trust in veterinary services and perceived disease severity that influence preventive practices.

**How this study might affect research, practice or policy:**

- The study findings advocate for culturally tailored health education that goes beyond information dissemination to address underlying beliefs and social structures.
- Policymakers, veterinary and public health practitioners can leverage the SEM model to target zoonotic disease control programs at the most influential behavioral drivers.
- To be effective future zoonotic disease control interventions must integrate traditional knowledge and local contexts into One Health frameworks to promote and sustain effective behavioral change.
- The study sets a precedent for integrating qualitative and quantitative methods in One Health research, promoting interdisciplinary collaboration.
- This integrated mixed-methods and structural modelling approach offers a replicable framework for studying zoonoses in other marginalized populations.

## INTRODUCTION

Brucellosis is a debilitating zoonotic disease caused by various bacteria of the genus *Brucella,* which affects both humans and a wide range of domestic animals, including cattle, goats, sheep, and camels (1,2). It is transmitted to humans through direct contact with infected animals and consumption of contaminated animal products. In low- and middle-income countries, particularly among pastoralist populations, brucellosis is endemic due to close human-livestock interactions, limited access to healthcare, and deep-rooted cultural practices that promote disease transmission (3,4). The daily proximity to livestock, through milking, assisting births, and handling of animal products, creates multiple pathways for zoonotic transmission. Globally, more than 2 million new human cases are reported annually (5).

Brucellosis results in enormous socioeconomic losses through reproductive failures, livestock abortions, reduced fertility, and decreased milk production, severely impacting agricultural productivity (6). Public health impacts of brucellosis include acute and chronic debilitating febrile illness with increased healthcare costs, reduced quality of life of affected individuals, and reduced productivity due to disability (7).

Kajiado and Marsabit counties in Kenya are predominantly inhabited by pastoralists, who maintain intricate livestock-based economies that are shaped by environmental, cultural, and social dynamics (8,9). Livestock-related practices such as the consumption of raw milk and meat, handling of skins and hides, lay treatment of sick animals, use of animal products in traditional medicine, and direct contact with aborted materials are culturally embedded and remain common despite their known association with transmission of zoonotic diseases such as brucellosis (10). Behavioral change initiatives frequently fail in these settings unless they are designed in close consultation with local communities in a way that accounts for their lived experiences and ecological constraints (11). Therefore, understanding how knowledge, practices, and cultural norms intersect to influence disease risk is critical for developing effective, context-specific One Health interventions.

The relationship between knowledge of brucellosis and subsequent behavior, attitudes, and safe practices is complex and often counterintuitive (12,13). Despite varying levels of awareness about brucellosis, studies indicate that knowledge alone does not significantly influence protective behaviors or attitudes towards the disease (14). In a study among Arab Israelis, 41% consumed non-pasteurized dairy products, despite high knowledge about brucellosis (15). Similarly, another study assessing brucellosis knowledge, attitudes and practice (KAP) among Arab Israelis reported that participants knowledge did not influence their practices despite 56.8% of them being aware of the disease (16). Research by Obonyo *et al.,* (2015) among pastoralist community in Garissa, Kenya, found that although 79% of pastoralists had heard of brucellosis, their attitudes and practices were poor (17). For instance, only 14% knew it could be prevented in animals and 46% indicated they would take no action even if they observed cases of livestock abortions in their herd.

Structural regression models such as Partial Least Squares Structural Equation Modeling (PLS-SEM) have been applied widely in KAP research within public health and livestock management systems to analyze the complex interrelationships between latent variables such as knowledge, attitude, and practice, particularly in settings where behavioral data are complex and sample sizes are modest. For instance, Usman *et al.*, (2023) utilized PLS-SEM to analyze the nexus between climate change, agriculture and food security among dairy farmers in Pakistan, revealing that knowledge significantly influenced preventive practices through the mediating role of attitudes, reinforcing the K→A→P pathway (18). Similarly, Chumsang *et al*., (2025) applied PLS-SEM in Thailand to assess livestock handlers’ KAP, demonstrating that formative indicators of knowledge (e.g., disease transmission routes) had stronger predictive power on safe practices than reflective attitude measures (19). These studies highlight PLS-SEM’s flexibility and effectiveness in uncovering latent behavioral constructs and estimating direct and indirect pathways, particularly the K → A → P structure commonly deployed in One Health research. Existing research on zoonotic disease transmission often focusses on epidemiological, biomedical and ecological dimensions (20–22), but rarely integrates socio-cultural perspectives with epidemiological assessments and structural path analyses.

Our study addresses this gap by examining sociocultural determinants and psychometric predictors (knowledge, attitude, practice) of brucellosis risk among pastoralist communities through a mixed-method and structural modelling approach. The study advances existing literature by refining practice-related indicators through exploratory factor analysis (EFA) and testing a hierarchical KAP model in a high-risk pastoralist context. Eventually, by highlighting how local context shapes both risk understanding and response behavior, this study aims to inform more equitable and effective approaches to zoonotic disease prevention in low-and middle-income pastoralist settings.

## MATERIALS AND METHODS

### Study sites

The study was conducted in Marsabit and Kajiado Counties in Kenya (Figure 1). Both counties are largely arid or semi-arid, with nomadic pastoral practices. Marsabit is located in northern Kenya and covers an area of 70,961.2 km^2^ with a population of 515,000 (23). The County has three main livelihood zones: pastoral (81%), agro-pastoral (16%), and others, including fishing, sandmining and businesses (3%) (8).

**Figure 1:**
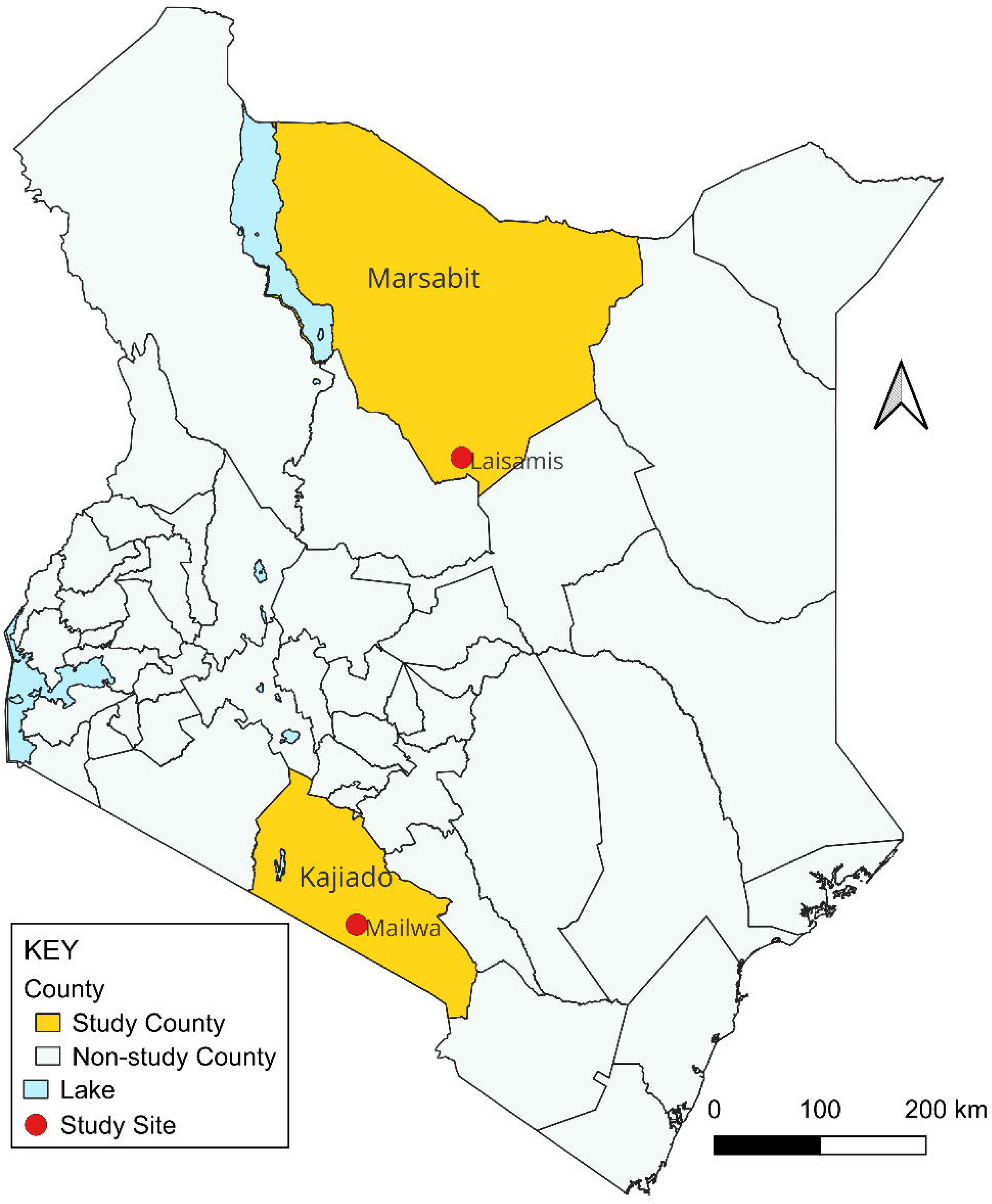
Map of Kenya showing the study sites. Yellow shows study counties (Marsabit County to the north and Kajiado County to the south); Red dots show location of the study sites. Source: Stella Mamuti

Kajiado County is located in the Rift Valley of southern Kenya. The county occupies 21,902 km^2^ with 1,268,261 people (23) and the estimated distribution by livelihoods as follows: 42% pastoralists, 35% in formal employment or casual labor, 12% agro-pastoralists, and 8% deriving their livelihood from mixed farming (9).

### Study Design

We employed convergent mixed-methods design combining household surveys, focus group discussion (FGDs), and individual in-depth interviews sequentially. A validated KAP scoring framework was implemented, and PLS-SEM approach used for path modelling. This multi-method approach provides invaluable insights often overlooked by singular quantitative or qualitative studies.

### Sample size and Sampling

This study was nested in a prospective cohort study that evaluated the role of camels and other livestock in the transmission of *Brucella* spp. and the Middle East Respiratory Syndrome Coronavirus in Kenya (Scientific and Ethics Review Unit reference # SERU 4405). The sample size was determined using Fleiss formula for cohort studies (24). Households were eligible for the study if they met the following criteria: they were located within 30 kilometers of the study health facilities (Laisamis Sub-County Hospital for Marsabit and Mailwa Health Centre for Kajiado), they raised at least one type of livestock species (such as cattle, camels, goats, or sheep), and the head of the household provided informed consent for their household and livestock to be included in the study. Human participants were eligible if they had resided in an eligible household for at least four months preceding the study and provided individual informed consent or assent to participate.

Participants for narrative interviews were selected purposively from enrolled household members while key informants included One Health practitioners and local leaders from the respective counties. Quantitative surveys were conducted among household heads (n=327) and adult household members. One adult per household was interviewed for practice-related questions (n=576), focus group discussions (FGDs) (n=20; 127 participants), individual in-depth interviews (IDIs) (n=68), key informant interviews (KIIs) (n=19), and informal participant observations and ‘go-along’ conversations (n=22).

For the household surveys on brucellosis-related awareness and practices, standardized questionnaires preloaded onto the REDCap mobile application, (Vanderbilt University Medical Center, Tennessee, USA), were administered electronically using password-protected tablets – with built-in data validation algorithms. These questionnaires collected data on socio-demographics, individual medical history, potential exposure to brucellosis, knowledge of the signs of brucellosis, history and frequency of contact with livestock, symptoms of acute brucellosis, health-seeking behaviors, and any treatment history within the previous 3 to 12 months.

In-depth individual interviews and focus group discussions were held to explore pastoralists’ knowledge and practices, including brucellosis manifestations and management strategies for prevention and treatment. Finally, key informant interviews were conducted with One Health practitioners and local leaders to capture knowledge and practices related to prevention and control of brucellosis. While quantitative methods can collect numerical data on pastoralists’ practices, they can’t get in depth information on why and how these practices emerge, spread and prevail.

### Patients and public involvement

Patients were not involved in this study. However, we conducted extensive participatory community engagement before study initiation and provided ongoing feedback throughout the study. We held initial entry forums with the county and subcounty One Health Unit leadership, elders, chiefs, *morans’* and women’s representatives to explain the study objectives, ensure cultural appropriateness of study procedures, enhance community acceptance, and address cultural sensitivities. We also held quarterly debriefs with the local administrators and community health workers and cascaded annual stakeholder forums to provide progress updates and address any new or emerging concerns. All study research assistants were recruited from the local community providing essential linguistic and cultural context during data collection, and ensuring mutual trust and data quality

### Construction of latent and reflective variables for brucellosis KAP assessment

Using syndromic terminology and pastoralists’ local knowledge of brucellosis signs and symptoms in animals and humans, we developed a composite KAP scoring framework using both binary (Yes/No) and multiple questions from the household enrolment survey. This scoring approach was adapted from methodologies in previous knowledge, attitude, and practice studies on zoonotic diseases (25,26) and refined by incorporating local epidemiological insights and behaviorally relevant variables. The modified algorithm included a broader set of knowledge and practice indicators such as knowledge of signs and symptoms of disease in animals and humans, knowledge of transmission routes, perceived disease seriousness, willingness to vaccinate, cull, and safely dispose of aborted materials (Appendix 1). This allowed a multidimensional evaluation of KAP, covering disease syndromes, transmission routes, prevention options, milk handling, and other sanitary practices.

Each binary question contributed one point for a “*Yes*” response and zero points for a “*No*” response, except P1, P2 and P6 where *“Yes”* and *“No”* responses scored zero and one, respectively. Various multiple-choice questions were incorporated to capture more detailed KAP predictor variables. Each selected option was assigned a score of one (Appendix 1). Each participant’s total score per latent variable sub-scale was obtained by summing their scores across all binary and multiple-choice items. This raw score was then normalized to a percentage by dividing it by the maximum attainable score. Pearson’s Chi-squared test or Fisher’s exact test (where applicable) was used to compare knowledge, attitude, and practice levels across sites for each variable.

### Application of PLS-SEM analysis to brucellosis KAPs

We employed Partial Least Squares Structural Equation Modeling (PLS-SEM) to analyze the causal relationship between Knowledge, Attitude, and Practice (KAP) on brucellosis among pastoralist communities in the study. PLS-SEM is a variance-based method optimized for predictive and exploratory modeling, particularly when dealing with non-normal data, formative indicators, or multiple latent constructs (27). PLS-SEM was selected due to its suitability for complex, multivariate behavioral data, its ability to handle latent (unobserved) constructs, and its robustness with modest sample sizes – 327 households in our case.

The study specified 23 questions in the household enrollment survey to measure brucellosis latent variables: Knowledge (9 items), Attitude (6 items), and Practice (8 items). The study was premised on the theoretical framework and null hypothesis that attitude towards brucellosis does not mediate the relationship between knowledge of brucellosis and the adoption of effective preventive practices among pastoral communities (i.e., the indirect effect of knowledge on preventive practices through attitude is not statistically significant; β = 0). This hierarchical approach aligns with behavioral theory (28) and empirical evidence suggesting that knowledge alone may not directly drive behavioral change but operates indirectly through attitudinal shifts (19).

### Data analysis and modelling procedures

Quantitative data were collected electronically using tablets through the REDCap mobile application (Vanderbilt University Medical Center, Tennessee, USA). The survey data were then loaded into and analyzed using R Software (R Foundation for Statistical Computing, Vienna, Austria). Descriptive and inferential statistics were carried out to summarize the sociodemographic characteristics of study participants, and explore the distribution and patterns of brucellosis knowledge, attitude and practices.

To further assess the causal relationship of the KAP constructs, PLS-SEM was applied using the *plspm* package in R software. This approach also facilitated the evaluation of the constructs’ reliability and validity and the causal pathways between knowledge, attitude and practices regarding brucellosis. The latent constructs (K, A, P) were modeled with reflective indicators. Exploratory factor analyses with reflective indicators were used to assess the sensitivity, robustness, reliability, and unidimensionality of the PLS-SEM model. Following exploratory factor analysis, we retained 5 indicators for Knowledge, 4 for Attitude, and 3 for Practice. Items were excluded if they failed to reach the pre-specified standardized unidimensionality or loading threshold of 0.50, ensuring that only reliable, conceptually coherent indicators contributed to each construct. Our structural expectations were that knowledge positively influences attitude, and that both knowledge and attitude promote desirable practice.

For the qualitative data, audio-recorded interviews were transcribed verbatim in Microsoft Word. Thematic analysis was performed using NVivo 14 (version 14.23.2, QSR International Pty Ltd. Lumivero, Denver, USA), to explore participants’ perceptions of brucellosis transmission, symptoms, treatment and prevention practices. The analysis followed Braun and Clarke’s framework (29), which includes familiarizing with the data, generating initial codes, searching for themes, reviewing themes, and presenting results. The process yielded codes on brucellosis manifestations in livestock and humans, causes, perceived severity, seasonality of the disease and treatment choices. The results were disaggregated by study site and presented in tables, figures, narrative descriptions and verbatim quotations.

#### Ethical considerations

The study protocol was reviewed and approved by the Kenya Medical Research Institute’s Scientific Ethics Review Unit (KEMRI-SERU) (reference #SERU 4405), and the KEMRI Animal Care and Use Committee (reference # KEMRI/ACUC/02.07.2022). A research permit was obtained from the National Commission for Science, Technology, and Innovations (License No: NACOSTI/P/22/17621). Permission to conduct the study was obtained from the Director General for Health, and the Director for Veterinary Services. The study conformed to the principles of the Helsinki Declaration. The researchers obtained written informed consent from all study participants before their engagement.

## RESULTS

### Socio-demographic characteristics

We compared key demographic, knowledge, attitude, and practice-related factors between respondents from Marsabit and Kajiado counties. Statistically significant differences were observed across several variables, indicating possible variations in brucellosis transmission risk and prevention awareness. The median age of respondents was 41 years (IQR: 33–51) in Kajiado and 43 years (IQR: 33–54) in Marsabit while the median household size was 7 (IQR: 5–8) in Kajiado compared to 6 (IQR: 4–7) in Marsabit (p = 0.002). A significantly higher proportion of respondents in Marsabit (96%) lacked formal education compared to only 71% in Kajiado, p-value *<*0.001. Most respondents from both counties identified as farmers or engaged in farm work as their primary occupation, as depicted in Table 1.

**Table 1:** Sociodemographic characteristics of the study participants in Kajiado and Marsabit Counties

| Characteristic | Kajiado<br>N = 153<sup>1</sup> | Marsabit<br>N = 174<sup>1</sup> | p-value |
| --- | --- | --- | --- |
| **Age (years), Median (IQR)** | 41 (33-51) | 43 (33-54) | 0.6 |
| **Age group** |  |  |  |
| 21 - 30 yrs | 22 (14%) | 18 (10%) | 0.3 |
| 31 - 40 yrs | 52 (34%) | 60 (34%) | >0.9 |
| 41 - 50 yrs | 34 (22%) | 41 (24%) | 0.8 |
| 51 - 60 yrs | 24 (16%) | 25 (14%) | 0.7 |
| 60 yrs + | 21 (14%) | 30 (17%) | 0.4 |
| **Sex of the household member** |  |  |  |
| Male | 106 (69%) | 87 (50%) | <0.001 |
| Female | 47 (31%) | 87 (50%) | <0.001 |
| **Highest level of education attained** |  |  |  |
| ^No formal education | 108 (71%) | 167 (96%) | <0.001 |
| Primary | 15 (9.8%) | 2 (1.1%) | <0.001 |
| Secondary | 17 (11%) | 2 (1.1%) | <0.001 |
| Post-secondary | 12 (7.8%) | 3 (1.7%) | 0.008 |
| Adult education | 1 (0.7%) | 0 (0%) | 0.5 |
| **Primary occupation** |  |  |  |
| Works on farm/Farmer | 120 (78%) | 130 (75%) | 0.4 |
| Housewife | 13 (8.5%) | 9 (5.2%) | 0.2 |
| Salaried off-farm skilled | 6 (3.9%) | 2 (1.1%) | 0.2 |
| Salaried off-farm non-skilled | 1 (0.7%) | 31 (18%) | <0.001 |
| Student | 1 (0.7%) | 1 (0.6%) | >0.9 |
| Other vocations | 12 (7.8%) | 1 (0.6%) | <0.001 |
| **Who usually makes decisions in the \*management of animals** |  |  |  |
| Household Head | 130(85%) | 160 (92%) | 0.047 |
| Spouse of Household Head | 21 (14%) | 11 (6.3%) | 0.025 |
| Son/Daughter | 1 (0.7%) | 3 (1.7%) | 0.6 |
| Father/Mother | 1 (0.7%) | 0 (0%) | 0.5 |

<sup>1</sup>No formal education means no school at all or attempted but did not complete primary school, \*Treatment, grazing, or sale of animals, IQR-interquartile range, Household head is an adult member of the household responsible for primary decision-making and livelihood of the household.

### Descriptive statistics of brucellosis knowledge, attitude, and practice (KAP) measurements

We assessed participants’ brucellosis-related knowledge, attitudes, and practices using a composite KAP scoring algorithm that integrated nuanced understanding of brucellosis through both binomial (Yes/No) questions and weighted multiple-choice responses. This allowed a multidimensional evaluation of KAP, covering disease syndromes, transmission routes, prevention options, milk handling, and other sanitary practices.

Overall, brucellosis knowledge, attitude, and practice levels were markedly low in both study sites. Notably, participants in Marsabit demonstrated significantly better knowledge (54% vs 36%, p-value <0.001), and attitude (45% vs 23%, p-value <0.001) but worse practice scores (26% vs 52%, p-value <0.001), respectively, compared to Kajiado – as shown in Table 2. In most households, 95% in Marsabit and 88% in Kajiado. have heard of brucellosis. Many also know that humans get infected by brucellosis (90% vs. 83%, p-value < 0.023), but few appreciate its impact in animals (46% vs. 39%, p-value < 0.135). Knowledge of specific aspects of brucellosis, such as signs and symptoms, or how it spreads in both humans and animals, was poor in both communities. Nevertheless, pastoralists in Marsabit manifested significantly better general brucellosis awareness, awareness of signs and symptoms in both humans and animals, and better awareness of its transmission in both humans and animals, respectively, compared to Kajiado. Furthermore, individuals in Marsabit were more aware of strategies to prevent brucellosis in humans (66% vs. 7%, p-value < 0.001) than in Kajiado county, while knowledge about protecting animals from contracting brucellosis was moderate in both counties (Table 2).

**Table 2:**
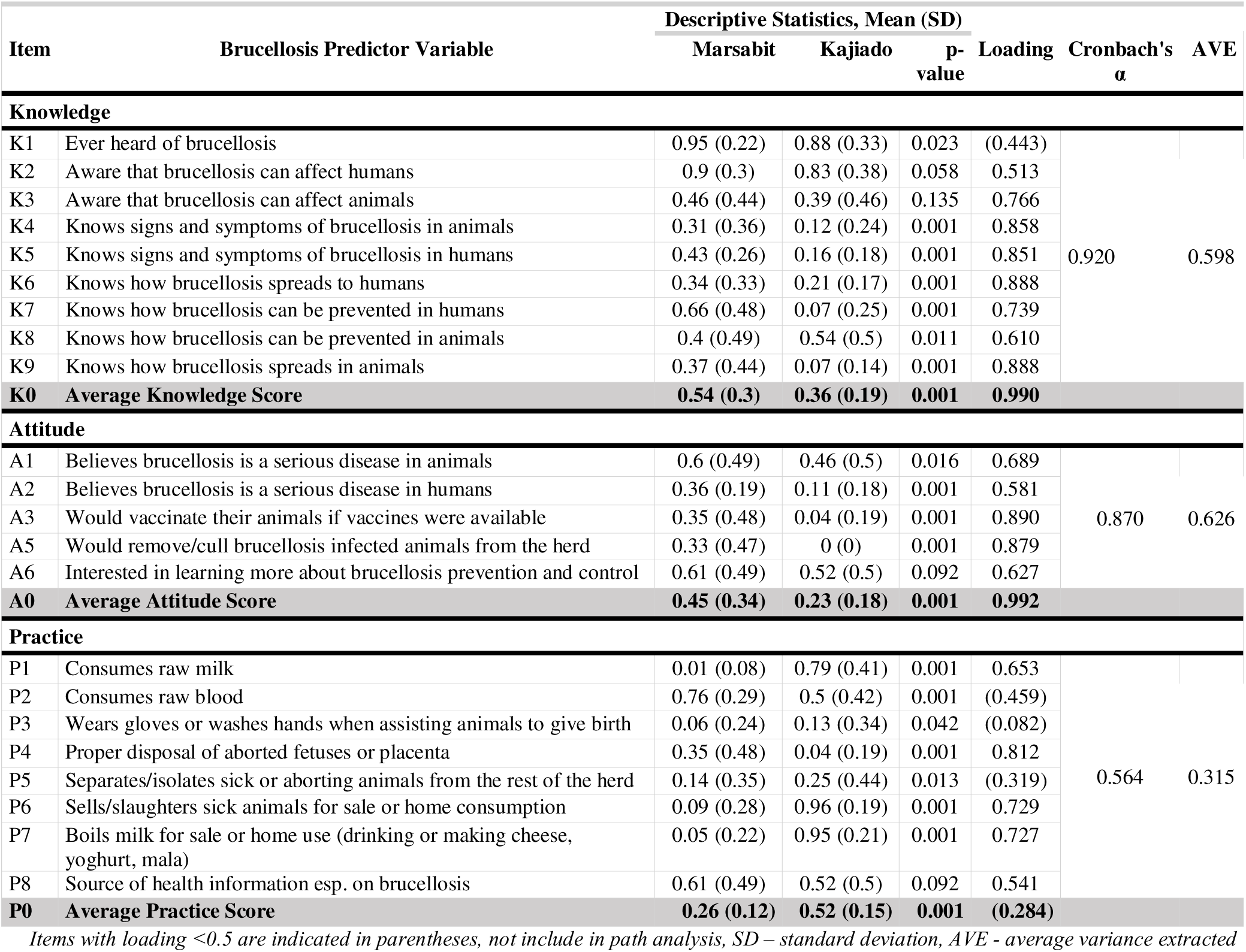
Brucellosis knowledge, attitude, and practice indicators by study site

### Pastoralist’s attitudes and beliefs

Attitudes toward brucellosis also differed across the counties, with pastoralists’ in Marsabit reporting relatively better appreciation of disease seriousness in humans (36% vs. 11%, p<0.001) and animals (60% vs. 46%, p=0.016), respectively, compared to Kajiado. They were also more likely to embrace preventive interventions like livestock vaccination (35% vs. 4%, p<0.001) or culling of infected animals (33% vs. 0%, p< 0.001), than households in Kajiado county (Table 2). Strikingly, none of the participants from Kajiado expressed willingness to isolate or cull brucellosis-infected animals. However, pastoralists from both sites were equally enthusiastic about gaining more knowledge on brucellosis – with 61% of participants from Marsabit and 52% from Kajiado expressing desire to learn more about brucellosis prevention and control, (p-value= 0.092).

### Pastoralist’s sanitary and dietary practices

While pastoralists in both sites recognized disease seriousness, practical commitment to control measures was low – 35% in Marsabit vs 4% in Kajiado. Despite reporting relatively worse knowledge and attitude scores, pastoralists in Kajiado demonstrated significantly better dietary and sanitary practices compared to Marsabit – with 95% vs 5% of households reportedly boiling milk before drinking, processing for other domestic use, or selling. On the other hand, risky behaviors such as drinking raw milk and blood were more prevalent in the Marsabit county (99% vs. 21%, p<0.001 and 76% vs. 50%, p< 0.001), respectively.

Milk treatment methods differed significantly between the sites. Boiling was nearly universally practiced in Kajiado (95%) but only rarely practiced in Marsabit, where 97% of individuals did not use any method to treat or preserve the milk. Other methods of milk preservation include medicinal herbs, ashes, and fermentation. While boiling milk is a proven preventive strategy against brucellosis, there exist cultural sanctions against boiling milk from animals.

> “Cultural norms dictate milk handling, with beliefs against boiling milk and a preference for raw milk consumption” (Male, IDI, #010, Marsabit).

With respect to preparation of homemade yoghurt, cheese and mala, striking differences were observed with all participants in Kajiado who reported making dairy products at home using boiled milk (52%), whereas in Marsabit, all 83 participants used unboiled milk, and none reported boiling milk, (*p<*0.001).

Consumption of uncooked blood was relatively common – 76% in Marsabit vs. 50% in Kajiado, (p< 0.001), suggesting an established cultural dietary practice. We explored this further through qualitative enquiry to ascertain the cultural veracity of this practice. Consumption of blood bore ritualistic value for the pastoralists as depicted by the following excerpts:

> “Blood is considered medicinal and consumed as part of meals, sometimes mixed with milk or porridge for sweetness and health benefits.” (Male, IDI, #039, Kajiado).
>
> “When you only use blood, it is not sweet, but when you add milk, it gets to be sweet, and you get satisfied after taking it.” (Female, IDI, #032, Marsabit).

Both communities have not embraced safe methods of disposing aborted animal materials or taking precautions during parturition assistance. Pastoralists in Kajiado demonstrated significantly worse disposal methods of placentae and aborted materials/fetuses (4% vs 35%, p<0.001), respectively. Various methods were used to dispose placentae and aborted materials in both counties, as highlighted in Figure 2. The most common method of disposing placentae and abortion materials was feeding them to dogs (96%), with significantly higher proportions in Marsabit (99%) compared to Kajiado (93%), p=0.01. Other common disposal practices included leaving the products of abortion or delivery in pasture (43%) or burying them in pit latrines (20%). Both practices were more prevalent in Marsabit compared to Kajiado (63% versus 20%, p<0.001 and 34% versus 3.3%, p<0.001), respectively.

**Figure 2:**
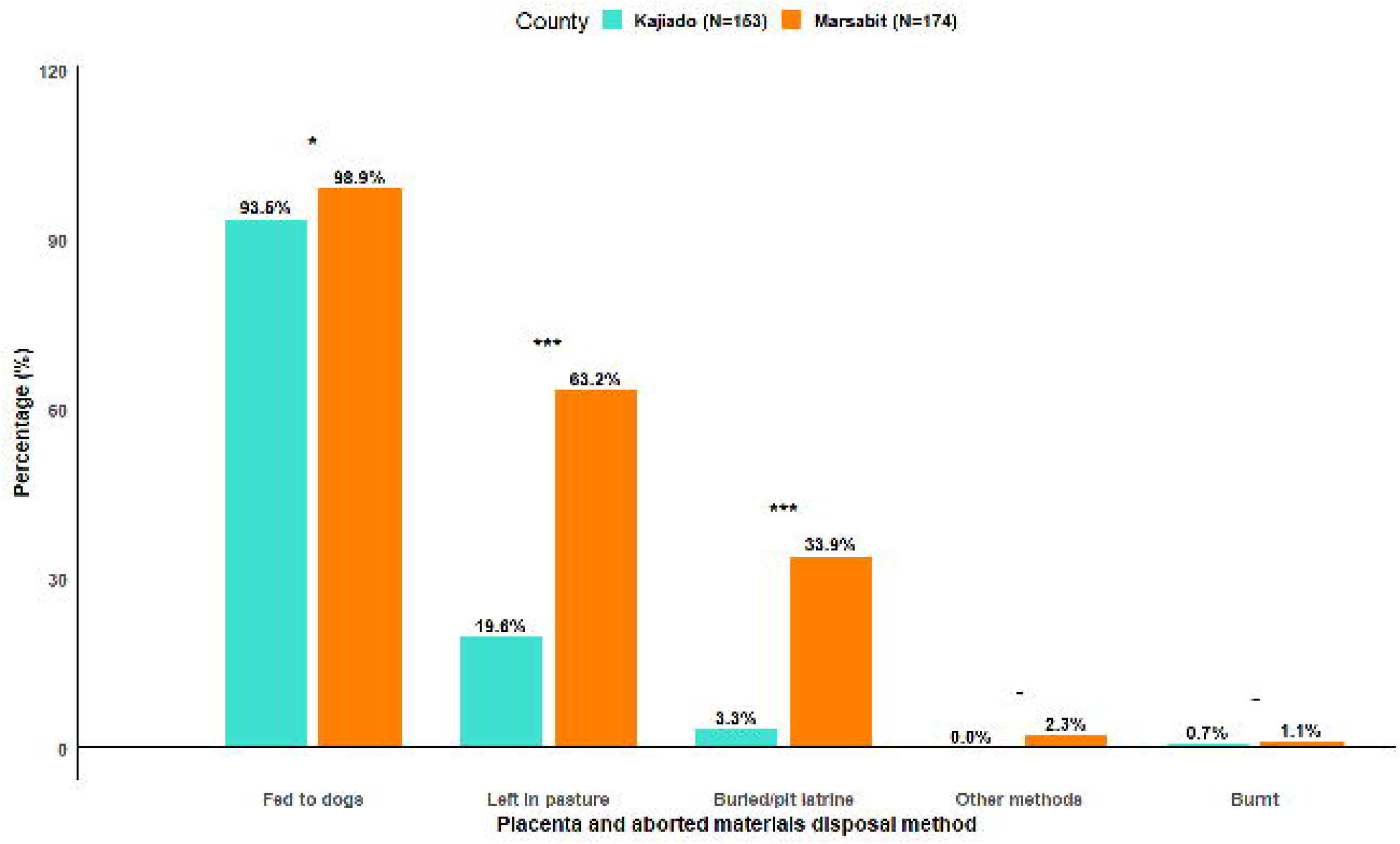
Comparison of disposal methods of placentae and aborted materials in Kajiado and Marsabit counties. *** means p < 0.001 (the observed difference between counties is highly significant, with less than a 0.1% probability of occurring by chance), ** means p < 0.01 (the difference is highly significant, with less than a 1% probability that it occurred by chance), * means p < 0.05 (there is a significant difference with less than a 5% chance that this difference occurred by random chance), - means there is no statistically meaningful difference between the two counties for that disposal method.

The survey results were corroborated by verbatim narratives from respondents during the ethnographic enquiry.

> “In case of miscarriage, the fetus is given to the dog.” (Male, IDI, #033, Kajiado).
>
> “Placenta is left for dogs if within the homestead, and for wild animals if outside.” (Female, IDI, #036, Kajiado).

Handling aborted fetuses with bare hands was common, given the near-universal practice of animal birth assistance within pastoral settings. The risk is worsened by the low transmission risk perception.

> “We just help them without gloves, for someone like me who is learned after helping them I go clean my hands, but then those from the reserves don’t wash their hands because they think the amniotic fluid has no problem, so they just wipe their hands on the bark of trees” (Male, IDI, #038, Marsabit).
>
> “Amniotic fluid can be used to help birth when water is not available.” (Male, IDI, #037, Marsabit).

### Handling of animals and animal products – manure, skins and hides

Regular handling of livestock was significantly higher in Kajiado across all species during the preceding 3 months, with sheep: 99% in Kajiado vs. 80% in Marsabit (p< 0.001), goats: 99% vs. 92% (p= 0.005), and cattle: 98% vs. 39% (p< 0.001), respectively (Table 3). Conversely, camels were exclusively reared in Marsabit, with 37% of respondents reporting regular contact with camels in the preceding 3 months. Slaughtering of wild animals was reported by 19% of respondents in Marsabit, compared to only 1.6% in Kajiado (p<0.001). High-risk activities such as slaughtering of wild animals, animal manure handling, and processing of fresh hides/skins were more frequently practiced in Marsabit compared to Kajiado.

**Table 3:** Handling of animals and animal products e.g. skins, hides, and manure

| Characteristic | Overall<br>n = 249<sup>1</sup> | Kajiado<br>n = 126<sup>1</sup> | Marsabit<br>n = 123<sup>1</sup> | p-value<sup>2</sup> |
| --- | --- | --- | --- | --- |
| Handling cattle on a regular basis in the past 3 months | 171 (69%) | 123 (98%) | 48 (39%) | <0.001 |
| Handling sheep on a regular basis in the past 3 months | 223 (90%) | 125 (99%) | 98 (80%) | <0.001 |
| Handling goats on a regular basis in the past 3 months | 238 (96%) | 125 (99%) | 113 (92%) | 0.005 |
| Handling camels on a regular basis in the past 3 months | 46 (18%) | 0 (0%) | 46 (37%) | <0.001 |
| Slaughtering of (wild) animals in the past 3 months | 25 (10%) | 2 (1.6%) | 23 (19%) | <0.001 |
| Handling/using animal waste in the past 3 months | 129 (52%) | 84 (67%) | 45 (37%) | <0.001 |
| Handling fresh animal hides/skins in the past 3 months | 70 (28%) | 27 (21%) | 43 (35%) | 0.018 |

<sup>1</sup>n (%); <sup>2</sup>Pearson's Chi-squared test; Fisher's exact test

### Reflective Measurement Model Assessment

An exploratory factor analysis (EFA) was initially conducted to assess the structure of the brucellosis KAP constructs. This was aimed at examining whether the observed variables clustered into the hypothesized latent construct. The reliability and validity of the initial model were evaluated using cross-loading and Average Variance Extracted (AVE). While most measurements met the recommended threshold of factor loading on the respective latent variables, several items showed low or weak cross-loadings, which indicates insufficient convergent validity. This refinement ensured that only measurement with acceptable psychometric attributes (i.e. a cross-loading threshold of 0.6 and their respective communalities exceeding 0.5), contributed to the estimation of the structural relationships between knowledge, attitude and practices of brucellosis. As shown in Table 2, the measurements with weak loadings, such as K1, K2, A2, P2, P3, P5, and P8, did not adequately describe the respective latent variable and were therefore not considered in the final PLS-SEM model.

Further, confirmatory factor analysis validated the internal consistency with Cronbach’s α values of 0.898 (Knowledge), 0.794 (Attitude), and 0.785 (Practice), all above the recommended 0.70 threshold. Composite reliability, measured using Dillon–Goldstein’s rho, was also high (0.926, 0.869, and 0.878, respectively). Convergent validity was confirmed, as the AVE exceeded 0.50 for all constructs (Knowledge = 0.717; Attitude = 0.627; Practice = 0.653). Mean communalities were consistent with the AVE values, further supporting indicator reliability.

The explanatory power of the structural model was strong, with R² = 0.848 for Attitude and R² = 0.572 for Practice, indicating that Knowledge had a substantial effect on Attitude and, together with Attitude, significantly explained Practice. The overall Goodness-of-Fit (GoF) index was 0.6901, demonstrating a satisfactory global model fit for the PLS-SEM framework (Table 4).

**Table 4:** Reliability and validity of the PSL-SEM model for the latent variables Knowledge,

| Latent Variable | Cronbach's Alpha<sup>2</sup> | Dillon-Goldstein's Rho<sup>2</sup> | AVE<sup>2</sup> | Mean Communality<sup>2</sup> | Mean Redundancy | R<sup>2</sup><sup>2</sup> |
| --- | --- | --- | --- | --- | --- | --- |
| Knowledge | 0.898 | 0.926 | 0.717 | 0.717 | 0.000 | 0.000 |
| Attitude | 0.794 | 0.869 | 0.627 | 0.627 | 0.531 | 0.848 |
| Practice | 0.785 | 0.878 | 0.653 | 0.653 | 0.374 | 0.572 |

<sup>1</sup>GoF (Goodness-of-Fit) = 0.6901; <sup>2</sup>Cronbach's Alpha and DG Rho assess reliability; AVE and Communality assess convergent validity; R<sup>2</sup> indicates explanatory power for endogenous variables.

The structural model explained substantial variance in both endogenous and exogenous constructs. As expected for an exogenous constructs in the model’s causal ordering and translation of measurement quality into predictive variance, Knowledge had near-zero redundancy, whereas Attitude (mean redundancy =0.531) and Practice (mean redundancy =0.374) showed substantive redundancy; given redundancy ≈ communality × R², these values imply variance of approximately 85% for Attitude (R² = 0.848) and 57% for Practice (R² = 0.572), as shown in Table 4.

The correlations among latent variables are positive and substantial, aligning with the hypothesized KAP structure: Knowledge correlates most strongly with Attitude (r = 0.902), followed by Attitude with Practice (r = 0.770) and Knowledge with Practice (r = 0.661), as shown in Table 8. This pattern anticipates the structural results in which Knowledge is closely associated with Attitude and, together, these constructs are linked to Practice; the relative magnitudes of the correlations mirror the strength of the corresponding paths reported in Table 5.

**Table 5:** Correlation between latent variables in the PLS-SEM Model

| Latent variables | Knowledge | Attitude | Practice |
| --- | --- | --- | --- |
| Knowledge | 1.000 | 0.902 | 0.661 |
| Attitude | 0.902 | 1.000 | 0.770 |
| Practice | 0.661 | 0.770 | 1.000 |

### Empirical Results of the PLS-SEM Model

The items in Figure 3 represent the measurements that were used to operationalize the latent constructs of knowledge, attitude and practices of brucellosis. The validated items demonstrated reliability, consistency, convergency validity and unidimensionality. Collectively, these measurement properties confirm that the items were suitable for assessing the causal pathways linking knowledge, attitude, and practices of brucellosis using structural equation modelling.

**Figure 3:**
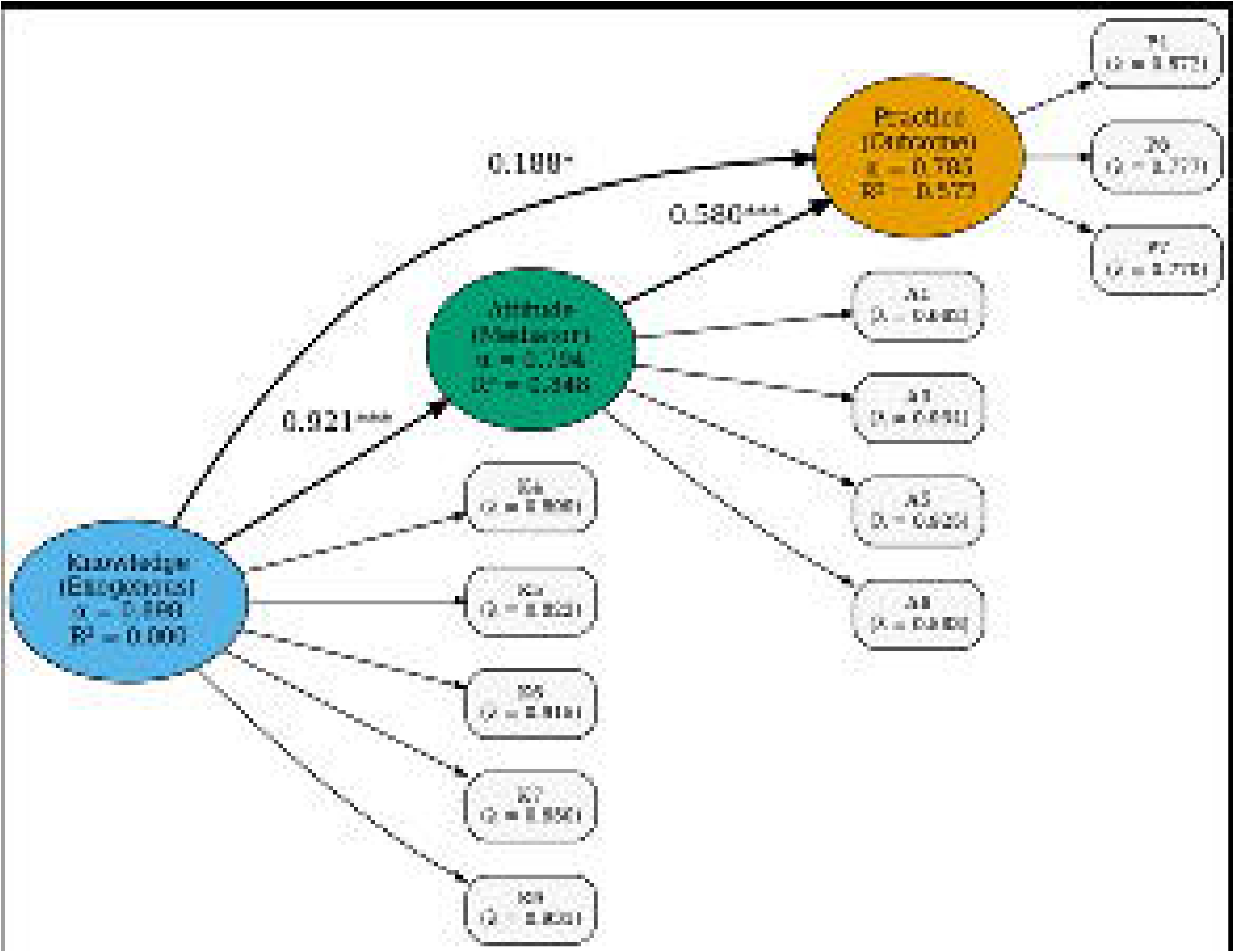
PLS-SEM of the Brucellosis Knowledge–Attitude–Practice framework. The model shows the direct and indirect causal relationship of knowledge on practice, mediated through attitudes. Standardized path coefficients are shown on factor loadings (λ) for each observed measurement. Reliability (Cronbach’s α) and adjusted R² are shown within each latent construct. The model realized a Goodness-of-Fit index of 0.6901, which is adequate to construct validity and model fit. Source: Authors own conception.

The structural equation model revealed that knowledge on brucellosis causes a strong and statistically significant effect on attitude (β = 0.921, p< 0.001), and attitude in turn significantly influences the adoption of preventive practices (β = 0.580, p< 0.001). Knowledge also had a modest but significant direct impact on practice (β = 0.188, p= 0.044). These findings demonstrate that higher level of knowledge about brucellosis is associated with more positive attitudes, which subsequently translates into improved adoption of preventive behaviors.

The indirect impact of knowledge on practice through attitude was substantial (β = 0.534), contributing most of the total effect (β = 0.722). This indicates that attitude is a critical mediating pathway between knowledge and behavior. Consequently, we reject the null hypothesis that attitude does not mediate the relationship between knowledge of brucellosis and adoption preventive practices. These findings underpin the importance of disseminating key messages that target the pastoralists’ attitude towards brucellosis for the communities to embrace effective prevention strategies.

## DISCUSSION

This study assessed and compared knowledge and practices related to brucellosis transmission among pastoralist communities in Kajiado and Marsabit counties in Kenya. We surveyed pastoral community members using validated questions for Knowledge (signs in animals; symptoms in people; transmission routes; prevention; spread among animals), Attitude (perceived seriousness; willingness to vaccinate; acceptance of culling; desire to learn more), and Practice (safe disposal of aborted materials; boiling milk; household milk treatment). The findings reveal significant inter-county differences in disease awareness, livestock handling practices, and socio-cultural determinants of exposure, underscoring the importance of tailoring public health and veterinary interventions to local contexts. A significantly higher number of female participants were enrolled in Marsabit, hence the low number of respondents regularly handling camels, as the Rendile culture and heritage prohibits women from handling camels. . The PLS model sought to evaluate the correlation between what people know about brucellosis, how they feel about it, and what they do in daily lives. Our measures were reliable, and the pattern is clear: knowledge shapes attitudes, and attitudes drive practice.

People showed solid understanding of how brucellosis spreads among animals and how humans get infected, while signs in animals and symptoms in people were less widely recognized. Knowledge about prevention in humans was mixed, suggesting room for clearer messaging on prevention and control interventions. Notably, although a significantly higher proportion of respondents in Marsabit (96%) lacked formal education compared to Kajiado (71%), their knowledge of brucellosis was significantly better than those in Kajiado (54% versus 36%). This disparity may reflect inequitable distribution of health infrastructure, and skewed penetration of public health interventions including exposure to disease messaging. It could also signify the fact that lack of formal education is not a barrier to acquisition and use of relevant knowledge. Previous research has shown that communities with better access to education and public health campaigns tend to have higher awareness of zoonotic diseases (12,30,31). The moderate-to-good knowledge scores observed in Marsabit may reflect higher baseline awareness, greater exposure to brucellosis communication, or localized outreach efforts by public health programs. Despite the remoteness and infrastructural challenges associated with the vast county of Marsabit, there are fairly well-established One Health networks coordinated by the county Government and supported by various humanitarian and non-governmental organizations such as PACIDA, USAID, CRS, and CONCERN worldwide that have been operating in the region for decades (32). At the core of this health promotion and disease surveillance system are community disease reporters (animal health), community health promoters (human health) and community draught reporters (environmental), which facilitates greater dissemination of health information, including brucellosis-related information. By contrast, continued reliance on traditional knowledge systems by most pastoralists in Kajiado may contribute to limited access to veterinary and public health education (33–36), and potentially expose them to health risks. Most respondents recognize brucellosis as serious and expressed desire to learn more about the disease. Openness to animal vaccination is promising, while acceptance of culling infected animals was understandably lower— this is a sensitive topic that likely needs respectful dialogue, compensation schemes, or alternatives (e.g., segregation) to gain support.

Cultural and occupational practices known to increase the risk of *Brucella* spp. transmission to humans were significantly more common in Marsabit compared to Kajiado. These include widespread consumption of raw milk, unprotected assistance during animal births, and handling sick or aborting livestock without protective gear or sanitation. There were clear regional differences in brucellosis-relevant risk practices between the two regions, with more frequent exposure to high-risk livestock activities and use of raw dairy products observed in Marsabit compared to Kajiado, where respondents were involved in relatively better dietary and hygiene practices. These behaviors and practices have previously been identified as high-risk in similar agro-pastoralist settings (2,37) and their continued occurrence suggests a gap in the uptake of protective health interventions and adoption of disease prevention practices. Furthermore, it suggests that even within broadly similar pastoralist systems, local knowledge systems, beliefs about disease causation, and access to services can create divergent pathways for zoonotic disease transmission. Slaughtering of wild animals poses a potential zoonotic bridge, particularly in areas with wildlife-livestock-human interfaces (38,39).

With respect to environmental contamination with potentially harmful pathogens such as brucellae, there was significantly more handling of animal manure/waste, hides and skins in Marsabit, suggesting higher environmental exposure to Brucella-contaminated materials. Poor placenta and abortus disposal methods, especially feeding dogs or leaving them in the pasture, were widespread, leading to increased environmental contamination potential and zoonotic disease transmission risk; as also observed in a study of bovine brucellosis in Pakistan, (40). Marsabit showed both better (more burial) and worse (more environmental exposure) disposal behaviors, likely reflecting mixed access to education and resources.

Although respondents in Kajiado practiced better milk hygiene practices compared to Marsabit where raw milk consumption is highly prevalent, the data suggests that they remain vulnerable due to limited milk sourcing diversity (41,42). Sourcing from neighbors or markets was significantly more common in Marsabit (p< 0.001), likely reflecting market access or cultural preferences. The consumption of raw milk, meat, and blood holds deep cultural significance, engrained in traditional practices, beliefs, and perceptions of the Maasai, Rendile and Samburu. Raw milk, sourced directly from cattle, stands as a staple food and a primary source of nourishment for both the young and old. It is revered for its nutritional value and esteemed for its role in traditional ceremonies and everyday sustenance. Similarly, raw meat, particularly from cattle, goats, or sheep, holds cultural importance, often consumed in ceremonies to mark significant events or as part of communal feasts, signifying unity and shared identity among the pastoral communities. Interventions on milk safety should therefore consider these context-specific behaviors to be effective. This practice not only provides sustenance but also embodies the pastoralist’s spiritual and cultural relationship with their animals, reflecting their reverence for nature and their pastoral way of life despite the zoonotic transmission risks. While such practices are often rooted in cultural norms and economic necessity, they highlight the complex interplay between knowledge, belief, and behavior in disease transmission dynamics (13).

Despite brucellosis being endemic in Kenya and designated as a notifiable zoonosis (20), our study results broadly indicate limited penetration of consistent and culturally resonant One Health messaging and preventive strategies into remote pastoralist regions. The findings also echo those of McDermott and Arimi (2002) who argued that disease control strategies must account for the livelihood realities of pastoralist populations, whose survival is closely tied to traditional livestock practices (10). Handling of cattle, sheep and goats was significantly more common in Kajiado compared to Marsabit. This potentially reflects the differing livestock production systems, with a more sedentary or mixed farming in Kajiado versus a more nomadic-pastoral and camel-heavy system in Marsabit. The frequent handling of livestock across both sites underscores the constant occupational exposure risks to *Brucella* spp., supporting the need for community targeted One Health interventions. In Marsabit, for instance, a predominant reliance on livestock for subsistence and livelihood, coupled with traditional herding practices, may entrench risky behaviors and normalize exposures that facilitate transmission. In this context, imposing behavior change without understanding local dynamics and constraints may have limited impact.

The PLS-SEM reveal a clear sequential pathway: knowledge about brucellosis among pastoralists in Kenya strongly shapes their attitudes, which in turn drive their practices. In our model, knowledge had a very large positive effect on attitude (β=0.921), and attitude likewise had a substantial effect on practice (β=0.580). Notably, the direct path from knowledge to practice was negligible, indicating full mediation by attitude. This finding aligns with the classic KAP paradigm, where awareness and understanding of a disease foster more favorable attitudes (e.g. risk perception, willingness to act), which then facilitate the adoption of safer practices. It is broadly consistent with reports from other low-resource settings: for instance, a rural Iranian study found that KAP levels on brucellosis were uniformly low, yet knowledge, attitude, and practice scores were significantly inter-correlated, suggesting a coherent KAP progression (26). Likewise, smallholder farmers in Pakistan exhibited pervasive knowledge gaps – 97% were unaware of brucellosis transmission modes – and nearly all engaged in at least one high-risk husbandry practice; those who had never heard of the disease or lacked formal education had significantly more risky behaviors (43). Such patterns underscore that inadequate knowledge often coincides with lax attitudes and unsafe practices, reinforcing the notion that improving knowledge is fundamental to changing mindsets and behaviors. Our model shows a strong path from Knowledge to Attitude and from Attitude to Practice. That means information alone isn’t adequate – people are more likely to change their behavior and practices when information shifts how they feel (risk, norms, efficacy). Hence, pairing education with attitude-shaping elements such as hands-on training, role models, behavior communication and change ambassadors, and community norms matter.

Our findings also have broader implications for zoonotic disease control in similar semi-arid regions of sub-Saharan Africa. They support the growing call for participatory, contextually embedded One Health strategies that leverage existing community structures, without undermining traditional knowledge and capacities to promote safe practices (44,45). As multiple studies have shown, disease control efforts that fail to consider socio-cultural and ecological realities tend to be ineffective or unsustainable (46,47). Leveraging local actors such as community animal and public health workers, elders, and livestock market influencers can strengthen the cultural relevance of health messages and promote greater community ownership of disease prevention efforts (11,48,49). This is particularly critical in arid and semi-arid regions, where formal veterinary and public health systems are often sparse or poorly coordinated.

## CONCLUSION

This study provides compelling insights into how socio-demographic factors, cultural practices, and knowledge systems influence brucellosis transmission risk among pastoralist populations in Kajiado and Marsabit counties. Given the zoonotic nature of brucellosis and its links to livestock handling and dairy consumption, the integration of veterinary and human health key messaging under a One Health framework may be essential to bridge these knowledge gaps. The substantial differences in awareness and exposure-related behavior between these counties highlight the need for geographically and culturally sensitive approaches to zoonotic disease prevention and control. Attitude plays a critical mediating role in the cause-and-effect pathway between knowledge and behavior. Consequently, One Health interventions for brucellosis control should emphasize disseminating knowledge that targets to change the pastoralists’ attitude towards brucellosis for the communities to embrace effective prevention strategies. Strengthening community engagement, improving access to veterinary and public health services, and integrating traditional knowledge holders into One Health initiatives will be key to reducing brucellosis transmission and improving pastoralist health outcomes in Kenya and beyond.

## Supporting information

Supplementary Table (Appendix) 1_(Proposed scoring framework for assessing brucellosis-related knowledge, attitudes, and practices (KAP)

## Data Availability

All data produced in the present study are available upon reasonable request to the authors.

## Supplementary Information

### Acknowledgements

We acknowledge the Kenya Medical Research Institute, the Ministry of Health, the Directorate of Veterinary Services, and the County Governments of Kajiado and Marsabit for their collaborative technical, administrative, and operational support in the implementation of the study. We are equally indebted to the farmers, their families and the pastoralist communities in Kajiado and Marsabit counties for invaluable insights on the operational design and participation in the study. Lastly, we immensely thank the study staff whose dedication and selflessness helped conduct the study to the highest standards: Nick Nalo, David Maina, Boru Wato, Millicent Minayo, Michael Baariu, Yusuf Galmogle, Liban Waqo, Moses, Learamo, Tonny Ngage, Asha Omar Abdikadir, Abdulahi Magarre, Moses Learote, Sylvester Leruk, Bowman Lewagat, Arthur Orieny, Peter Otom, Melita Mark Lein, Raphael Pasha, Benson Mutete, Kelvin Mukhwana, and Ntaisi Pore.

### Authors’ contributions

Conceptualization: DCO, SWM, DO, MKN, EO

Funding acquisition: WJ, MKN, EO

Methodology: DCO, SWM, DO, MKN, EO

Investigation: DCO, SWM, DO, RN, BB, NWN, JMN, SAK, EO

Project administration: DCO, RN, SAK, MKN, EO

Supervision: DCO, SWM, DO, RN, JG, SAK, WJ, MKN, EO

Data curation: DCO, SWM, DO, BO, RN, EO

Formal analysis: DCO, SWM, DO, BO, JAK, EO

Resources/Software: DO, SLN, WJ, MKN, EO

Validation: DCO, SWM, DO, BO, JAK, JG, SLN, EO

Visualization: DCO, SWM, DO, BO, EO

Writing - original draft: DCO, SWM, DO, BO

Review and editing: DCO, SWM, DO, BO, RN, JAK, BB, JG, SLN, NWN, JMN, SAK, HKN, EK, WJ, MKN, EO

All authors have read and agreed to the published version of the manuscript.

### Funding

This work was supported by the U.S. Department of Defense’s Defense Threat Reduction Agency under DTRA GRANT#12988984, and the U.S. National Institutes of Health-Institute of Allergy and Infectious Diseases under Award Number D43TW011519. The funders had no role in study design, data collection and analysis, decision to publish, or preparation of the manuscript. The content of this report is solely the responsibility of the authors and does not necessarily represent the official views of NIH or DTRA.

### Declarations

#### Ethics approval and consent to participate

The study protocol and informed consent forms were reviewed and approved by the Kenya Medical Research Institute’s Scientific and Ethics Review Unit (KEMRI-SERU, reference # SERU 4405).

#### Informed Consent Statement

Written informed consent was obtained from all participants involved in the study.

#### Consent to publish

The authors declare consent for publication of this article in the BMJ Global Health journal.

#### Availability of data and materials

The study data is available at the Washington State University database and will be shared upon formal requests.

### List of abbreviations

ACUC: Animal Care and Use Committee
ASAL: Arid and Semi-arid Lands
AVE: Average Variance Extracted
CRS: Catholic Relief Services
DTRA: Defense Threat Reduction Agency
EFA: Exploratory Factor Analysis
FGD: Focus Group Discussion
GoF: Goodness-of-Fit
IDI: Individual In-depth Interview
IPC: Infection Prevention and Control
IQR: Inter-Quartile Range
KAP: Knowledge, Attitude, and Practices
KEMRI: Kenya Medical Research Institute
KII: Key Informant Interview
KNBS: Kenya National Bureau of Statistics
IPC: Infection Prevention and Control
KEMRI: Kenya Medical Research Institute
MoH: Ministry of Health
NACOSTI: National Commission for Science, Technology and Innovation
PLS-SEM: Partial Least Squares Structural Equation Modeling
REDCap: Research Electronic Data Capture
WSU GH: Washington State University Global Health
SERU: Scientific and Ethics Review Unit
Spp.: Species
R: R Statistical Software

## Appendix 1 Proposed scoring framework for assessing knowledge, attitudes, and practices (KAP) related to brucellosis, outlining item domains, response options, and corresponding weights applied to generate composite scores for each construct

