## Supplementary Table (Appendix) 1_(Proposed scoring framework for assessing brucellosis-related knowledge, attitudes, and practices (KAP) for "Integrating Ethnography and Structural Equation Modelling to Assess Brucellosis Knowledge, Attitude, and Practices among Pastoralist Communities in Kenya"

|  | *Appendix 1: Proposed scoring framework for assessing knowledge, attitudes, and practices (KAP) related to brucellosis, outlining item domains, response options, and corresponding weights applied to generate composite scores for each construct.* | | | |
| --- | --- | --- | --- | --- |
|  | Model item/measurement |  | Response | Score |
| Knowledge | **K1. Ever heard of brucellosis** |  | Yes (1), No (0) | 1.0 |
|  | **K2. Aware that brucellosis can affect humans** |  | Yes (1), No (0) | 1.0 |
|  | **K3. Aware that brucellosis can affect animals** |  | Yes (1), No (0) | 1.0 |
|  | **K4. Knows signs and symptoms of brucellosis in animals***. K_41_. Abortion K_42_. Weak offspring or still born K_43_. Retained placenta K_44_. Swollen testes* 7.0  *K_45_. Infertility/Prolonged calving interval K_46_. Reduced milk production K_47_. Joint swelling/lameness/hygromas* | | | |
|  | **K5. Knows signs and symptoms of brucellosis in humans** *K_51_. Undulant fever K_52_. Sweating K_53_. Chills K_54_. Headache K_55_. Fatigue / weakness / malaise* 8.0  *K_56_. Night sweats K_57_. Joint / muscle pain K_58_. Loss of appetite* | | | |
|  | **K6. Knows how brucellosis spreads to humans** *K_61_. Contact with aborted fetuses or birth materials K_62_. Assisting in animal birthing* 8.0  *K_63_. Contact with sick animals K_64_. Ingestion of raw milk K_65_. Eating raw/undercooked meat or blood*  *K_66._ Slaughtering/butchering K_67_. Handling skins/hides from infected animals K_68_. Inhalation of droplets* | | | |
|  | **K7. Knows how brucellosis can be prevented in humans** |  | Yes (1), No (0) | 1.0 |
|  | **K8. Knows how brucellosis can be prevented in animals** |  | Yes (1), No (0) | 1.0 |
|  | **K9. Knows how brucellosis spreads in animals** *K_91_. Ingestion of contaminated pasture K_92_. Ingestion of contaminated animal tissues* 5.0  *K_93_. Contact with sick animals/animal tissues K_94_. Drinking/sharing contaminated water*  *K_95_. Contact with wild animals* | | | |
|  |  |  |  | **33.0** |
| Attitude | **A1. Believes brucellosis is a serious disease in animals** |  | Yes (1), No (0) | 1.0 |
|  | **A2. Believes brucellosis is a serious disease in humans** |  | Yes (1), No (0) | 1.0 |
|  | **A3. Would vaccinate their animals if vaccines were available** |  | Yes (1), No (0) | 1.0 |
|  | **A4. Would report to the animal health worker if their animals aborted or looked ill** |  | Yes (1), No (0) | 1.0 |
|  | **A5. Would remove/cull Brucellosis infected animals from the herd** |  | Yes (1), No (0) | 1.0 |
|  | **A6.** **Interested in learning more about brucellosis prevention and control** |  | Yes (1), No (0) | 1.0 |
|  |  |  |  | **6.0** |
| Practices | **P1. Consumes raw milk** Yes (0), No (1) 1.0 | | | |
|  | **P2. Consumes raw blood** |  | Yes (0), No (1) | 1.0 |
|  | **P3. Wears gloves or washes hands when assisting animals to give birth** |  | Yes (1), No (0) | 1.0 |
|  | **P4.** **Proper disposal of aborted foetuses or placenta** *P_41_. Bury/throw in pit latrine P_42_. Burn* 2.0 | | | |
|  | **P5. Separates/isolates sick or aborting animals from the rest of the herd** |  | Yes (1), No (0) | 1.0 |
|  | **P6. Sells/slaughters sick animals for sale or home consumption** |  | Yes (0), No (1) | 1.0 |
|  | **P7. Boils milk for sale or home use (drinking, making yoghurt, cheese or mala)** |  | Yes (1), No (0) | 1.0 |
|  | **P8.** **Source of health information esp. on brucellosis** *P_81_. Health worker (human or veterinary) P_82_. Mass media (TV/radio/social media)* 6.0  *P_83_. Community gatherings/meetings P_84_. Religious leader P_85_. Family member/friend/neighbour P_86_. Poster/flyers* | | | |
|  |  |  |  | **14.0** |

*Knowledge sub-scale: 9 items: Total score=33. Poor knowledge = 0-11, average knowledge = 12-22, Good knowledge = 23-33. Attitude has 6 items: Total score=6. Poor attitude = 0-2, Fair attitude = 3-4, Good attitude = 5-6. Practice sub-scale has 8 items: Total score=14. Poor practice = 0-4, Fair practice = 5-9, Good/satisfactory practice = 10-14*
